# Prediction of plasma volume and total hemoglobin mass with machine learning

**DOI:** 10.1101/2023.02.17.23286080

**Authors:** B. Moreillon, B. Krumm, J.J. Saugy, M. Saugy, F. Botrè, J.-M. Vesin, R. Faiss

**Affiliations:** Union Cycliste International, World Cycling Centre, Aigle, Switzerland; Research and Expertise in anti-Doping Sciences (REDs), Institute of Sport Sciences, University of Lausanne, Lausanne, Switzerland; Laboratorio Antidoping, Federazione Medico Sportiva Italiana, Rome, Italy; Signal Processing Laboratory 2, Swiss Federal Institute of Technology, Lausanne, Switzerland

**Author notes:** Corresponding author: Raphael Faiss, PhD, PD, REDs, Synathlon Quartier Centre, Office 2316, 1015 Lausanne.

**Keywords:** Blood, Total hemoglobin mass, Plasma volume, Prediction, Machine learning

## Abstract

Anemia is a widespread disease commonly diagnosed through hemoglobin concentration ([Hb]) thresholds set by the World Health Organization (WHO). However, [Hb] is subject to significant variations mainly due to shifts in plasma volume (PV) which impair the diagnosis of anemia and other medical conditions. The aim of this study was to develop a model able to accurately predict total hemoglobin mass (Hbmass) and PV based on anthropometric and complete blood count (CBC) analyses. 769 CBC coupled to measures of Hbmass and PV using the CO-rebreathing method were used with a machine learning tool in a numeric computing platform (MATLAB regression learner app) to calculate the model. For the predicted values, root mean square error (RMSE) was of 37.9 g and 50.0 g for Hbmass, and 194 ml and 268 ml for PV, in women and men, respectively. Measured and predicted data were significantly correlated (p<0.001) with the coefficient of determination (R^2^) ranging from 0.73 to 0.81 for Hbmass, and PV, in both women and men. The bland-altman bias between estimated and measured variables was in average of -0.69 for Hbmass and 0.73 for PV. This study proposes a valid model with a high prediction potential for Hbmass and PV, providing relevant complementary data in numerous contexts. This method can notably bring information applicable to the epidemiology of anemia, particularly in countries with high prevalence or in specific population such as high-altitude communities.

## Introduction

Hemoglobin (Hb) is an essential iron-containing protein present in red blood cells (RBC). It binds to oxygen in the respiratory organs and is then transported in blood to body tissues where the oxygen is released to permit cellular respiration. Hemoglobin synthesis occurs in the erythrocytes and is divided in two main components: heme synthesis and globin synthesis. Heme synthesis takes place in both the cytosol and the mitochondria of the erythrocytes whereas globin synthesis only occurs in the cytosol. (1, 2). Hemoglobin levels in the body are usually reported in concentration (g/L or g/dL) as RBC are suspended in plasma and are mostly measured as part of a complete blood count (CBC). CBC is a rapid and relatively inexpensive method that offers important information regarding the diagnosis of different pathologies or conditions. Notably CBC, and [Hb], can be used to detect athletes using blood doping to increase their performance (3, 4), to identify individuals at risk of stroke in vulnerable populations (5, 6), to help patients hospitalization decision (7) or to regulate patient blood management (PBM) (8-10). But most importantly, CBC is used to diagnose hemoglobin disorders such as anemia. Anemia is a condition defined by [Hb] under certain reference ranges and can be due to a variety of causes such as nutritional deficiencies, inflammations, infections, or genetic conditions. The World Health Organization (WHO) established thresholds from 11 g/dL to 13 g/dL (depending on age and sex) for its diagnosis (11) and its prevalence can raise up to 80% depending on the population (12). Thus, early diagnosis is important to prevent chronic impairments (13-15).

However, numerous studies demonstrated the pre-analytical, analytical, and physiologic variability of [Hb] measurement (16, 17). It is highly dependent on plasma volume (PV) which is regulated by the renin-angiotensin-aldosterone system and is prone to significant fluctuations. Indeed, several studies demonstrated changes in PV in a variety of conditions such as heat, altitude, acute and chronic physical activity or hydration status (18-20). In this context, the information provided by CBC and [Hb], while robust, is still difficult to interpret for experts (21).

While [Hb] is highly dependent on PV and its variability, total hemoglobin mass (Hbmass) is a relatively stable parameter. It represents the absolute mass of circulating Hb in the body and several studies showed its stability in different situations, notably during altitude training camps or with exercise protocols (22-26). In the context of medical diagnosis, Hbmass measurement provides complementary and relevant information to the CBC. Current gold-standard methods to measure Hbmass are the infusion of radioactive isotope tracers such as sodium pertechnetate and sodium radiochromate (27, 28) or the use of fluorescent dyes such as indocyanine green (29). More recently the carbon monoxide (CO) rebreathing method has been developed and scientifically approved. Briefly, it consists of breathing during a brief period (2-10 min) a small given amount of CO, which has a much greater affinity to hemoglobin compared to oxygen, mixed with O^2^. By assessing the increase in carboxyhemoglobin in pre- and post-conditions, Hbmass and PV can be quantified (30-32). However, this method is rather time-consuming and more expensive than CBC. Moreover, it raises some practical and safety challenges in medical patients with conditions such as heart failure or coronary artery disease (33-36).

CBC is thus, a rapid and robust method that gives relevant information for the diagnosis of numerous medical conditions, but which is subject to potential high variability due to PV shifts. Hbmass measurement offers valuable complementary information but is more challenging. Therefore, the aim of this study was to design a computational model to estimate Hbmass and PV from the results of a simple CBC. We hypothesized that measured and predicted values would be in good agreement.

## Material and methods

### Study subjects

769 data points with concomitant CBC and PV measurements were gathered from different studies in which participants volunteered to take part. Briefly populations consisted of (1) men and women control subjects (n=44), (2) free-divers (n=15), (3) elite athletes (n=31). In total, 330 observations for women and 439 observations for men were collected. Procedures and risks were fully explained to the subjects, and all of them gave their written consent to participate in the study. All studies were approved by the local ethics committee (CER-VD, Lausanne, Switzerland) and conducted in accordance with the Declaration of Helsinki.

### Blood sampling

Blood sampling was conducted by two experienced phlebotomists following the current WADA guidelines on analytical procedures (37). The participants reported to the lab having avoided any physical exercise in the 2 hours preceding the sampling. The subjects then remained seated for 10 min to avoid acute PV variation (38). Whole blood was collected either in 2 mL tubes (BD Vacutainer® tubes (EDTA-K2 (K2) CE cat no. 368856/ref US 367856, BD Europe, Eysins, Switzerland) or 2.7 mL tubes (K2 EDTA 2.7 mL, Sarstedt AG, Nümbrecht, Germany) considered equivalent which were homogenised at room temperature on a roller for 15–45 min and then analysed in duplicate with a gold standard flow cytometry blood analyser (Sysmex XN-1000, Sysmex, Norderstedt, Germany). Internal quality controls provided by the manufacturer (Sysmex E-Checks, levels 2 and 3) were run twice before each batch of samples. The following variables were analysed: White Blood Cell count (WBC#), RBC, [Hb], Hematocrit (HCT), Mean Corpuscular Volume (MCV), Mean Corpuscular Hemoglobin (MCH), Mean Corpuscular Hemoglobin Concentration (MCHC), Platelets (PLT), Red Blood Cell Distribution Width (standard distribution) (RDW-SD), Red Blood Cell Distribution Width (coefficient of variation) (RDW-CV), Platelet distribution width (PDW), Mean platelets volume (MPV), Platelet-large cell ratio (P-LCR), Procalcitonin (PCT), Neutrophils count (NEUT#), Lymphocytes count (LYMPH#), Monocytes count (MONO#), Eosinophils count (EO#), Basophiles count (BASO#), Neutrophils percentage (NEUT%), Lymphocytes percentage (LYMPH%), Monocytes percentage (MONO%), Eosinophils percentage (EO%), Basophiles percentage (BASO%), Immunoglobulins count (IG#), Immunoglobulins percentage (IG%), Reticulocytes percentage (RET%), Reticulocytes count (RET#) and Immature Reticulocyte Fraction (IRF).

### Plasma volume and total hemoglobin mass measurement

A fully automated system was used to determine PV and HBmass using a CO rebreathing procedure (OpCo: Detalo Instruments, Birkerod, Denmark), which is outlined in detail elsewhere (31). Briefly, subjects were asked to breathe a 100% oxygen (O^2^) bolus for 1 min in order to flush the airways of nitrogen before a bolus of 1 mL/kg of 99.997% chemically pure CO (Carbagas, Liebefeld, Switzerland) was administered in the circuit and rebreathed for 9 min in a supine position. Venous blood samples of 1.2 mL (S-Monovette Li-Heparin, Sarstedt, Nümbrecht, Germany) were taken before and after the rebreathing phase for the determination of carboxyhemoglobin (HbCO%) in triplicate with a separate gasometer (ABL80 Co-Ox, Radiometer, Copenhagen, Denmark) for HBmass determination using formulas described elsewhere (31). The CO remaining in the system was measured with a CO meter (Monoxor Plus, Bacharach, New Kensington, Pennsylvania, USA) and subtracted from the initial amount introduced to define the exact CO bolus received with a 0.1 mL typical error (TE). The total red blood cell volume (RBCV), PV, and blood volume (BV) were then additionally calculated from HBmass, [Hb], and HCT. In this study, the TE for the determination of HBmass was 1.8%, which is in line with existing studies (31).

### MATLAB script

The models were calculated using MATLAB regression learner app (MATLAB R2022b, MathWorks, Inc, Natick, USA). Anthropometric (age (y), height (cm), and weight (kg)) and CBC (listed above) data were compiled and imported into MATLAB to be used as variables to predict PV and Hbmass. Data was discriminated depending on sex and the rational quadratic model from the Gaussian Process regression (GPR) was used. 4 different models were created to predict the following variables: Hbmass women, Hbmass men, PV women and PV men. An example session is displayed in figure 1.

**Figure 1:**
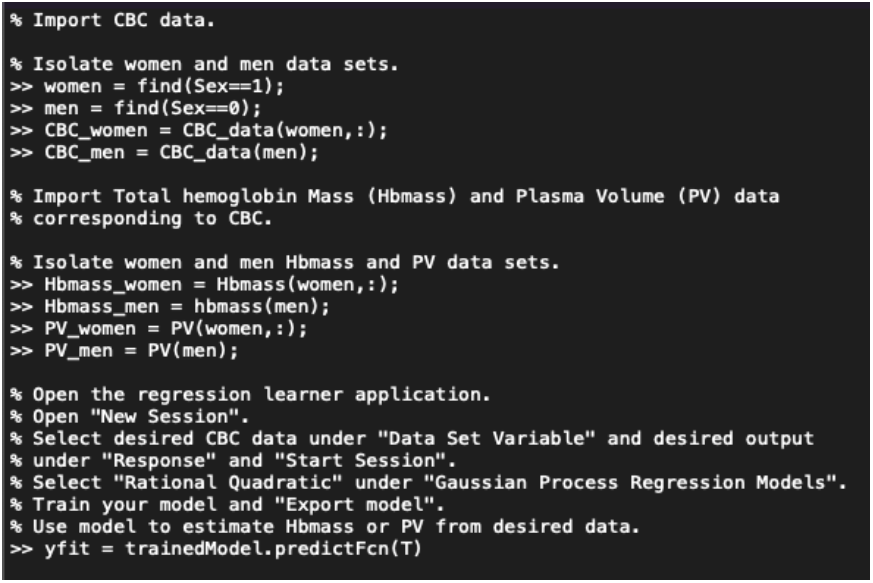
example of a Matlab session used to calculate predictor models

### Statistical analyses

Data are presented as means ± standard deviations (SD). To assess agreement between measured and predicted values of Hbmass and PV, root mean square error (RMSE), mean square error (MSE) and mean absolute error (MAE) were calculated. Additionally, Pearson’ s correlations and Bland-Altman analyses were performed. All statistical analyses were performed with dedicated softwares (MATLAB R2022b, MathWorks, Inc, Natick, USA; Prism, Version 8.4.2, GraphPad Software, La Jolla, California, USA).

## Results

Measured Hbmass was 610 ± 82 g for women, and 968 ± 114 g for men, Measured PV was 2987 ± 1461 ml for women, and 3774 ± 543 ml for men. In comparison with estimated values (i.e., RMSE), relative errors were respectively 6.1%, 6.3%, 5.2% and 7.2%. MAE, which tends to be less sensitive to outliers, is lower than RMSE for every parameter (Table 1). Measured and predicted values for Hbmass and PV were significantly correlated (p<0.001), and R^2^ were 0.79, 0.81, 0.74 and 0.75 for Hbmass women, Hbmass men, PV women and PV men, respectively (Figure 3). Finally, Bland-Altman’ s analyses showed that both methods are in good agreement with biases of 0.27, -1.66, 0.08 and 1.38 for Hbmass women, Hbmass men, PV women and PV men, respectively (Figure 4).

**Table 1:** Root Mean Square Error (RMSE), coefficient of determination (R^2^), Mean Square Error (MSE) and Mean Absolute Error (MAE) for total hemoglobin mass (Hbmass, g) and plasma volume (PV, ml) predictions in men and women.

|  |  | PV | Hbmass |
| --- | --- | --- | --- |
| Men | RMSE | 272 | 50.3 |
|  | MSE | 74254 | 2531 |
|  | MAE | 197 | 39.5 |
| Women | RMSE | 187 | 37.0 |
|  | MSE | 34941 | 1372 |
|  | MAE | 146 | 28.8 |

**Figure 2:**
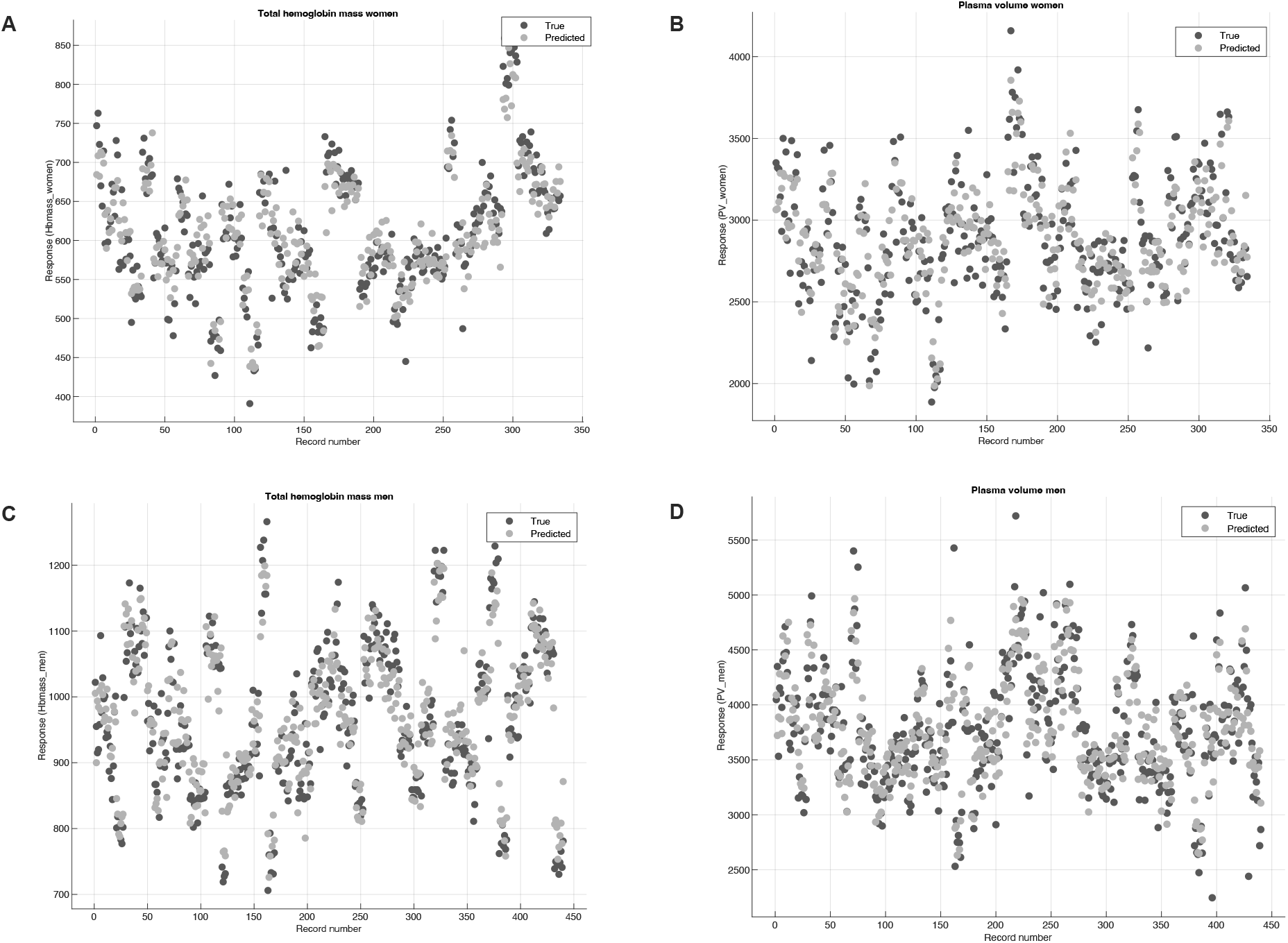
graphical representations of true (black) versus predicted (grey) total hemoglobin mass (Hbmass) and plasma volume (PV) for women (A, B, respectively) and men (C, D, respectively).

**Figure 3:**
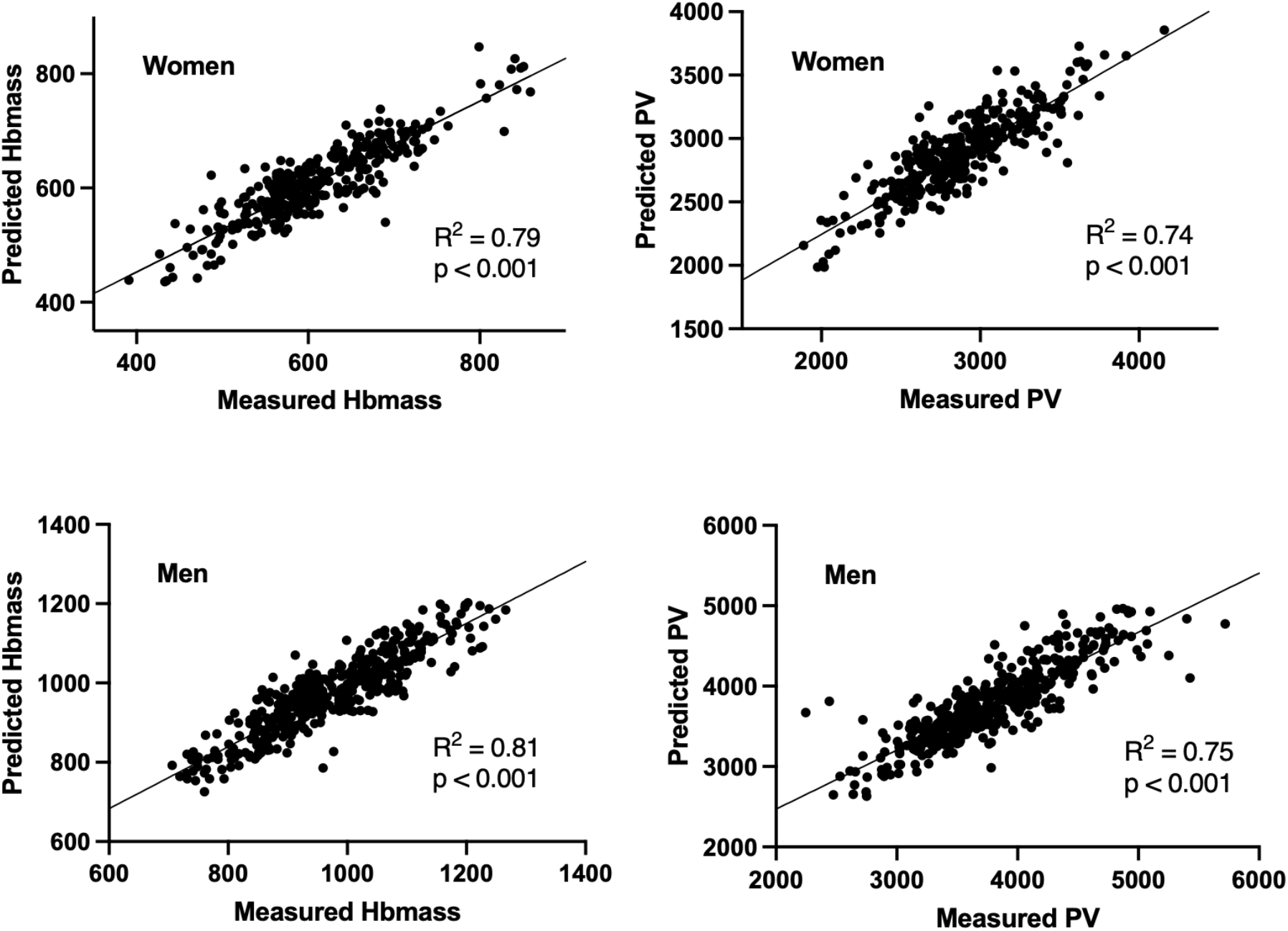
Correlations between measured and predicted total hemoglobin (Hbmass) (left) and plasma volume (PV) (right) for women and men, respectively.

**Figure 4:**
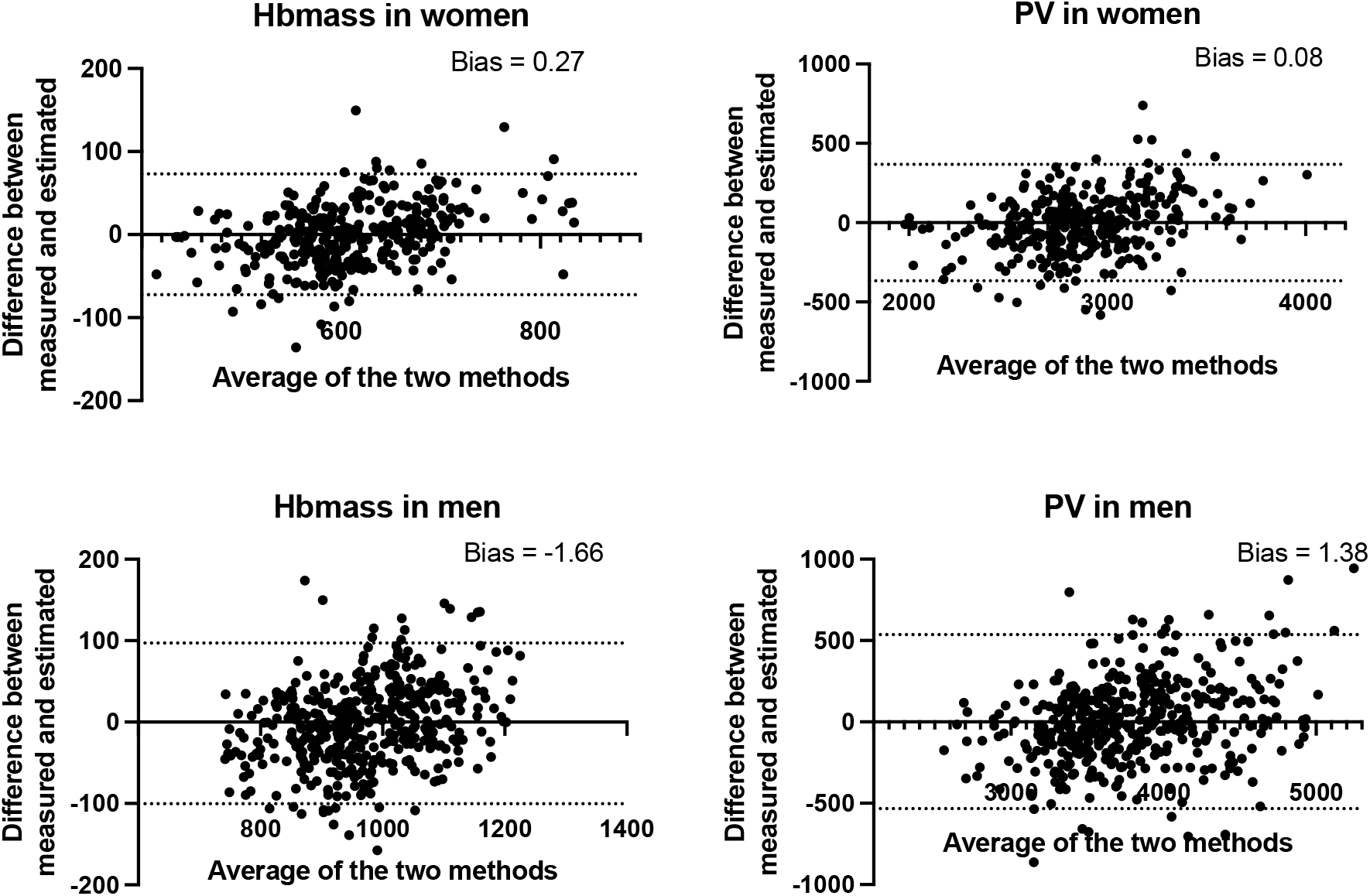
Bland–Altman plots comparing measured and predicted total hemoglobin (Hbmass) and plasma volume (PV) for women and men respectively. The difference is represented as function of the average values with 95% limits of agreement (dotted lines), computed as the mean difference (bias) plus or minus 1.96 times its SD

## Discussion

Hbmass and PV were accurately predicted for both women and men based on anthropometric and CBC analyses using a model calculated with MATLAB’ s regression learner application. Such a model can be of significant interest in a wide range of applications. Given the low relative RMSE of 6.1% for women and 5.2% for men in predicting Hbmass, this model adds valuable information to the simple CBC in many contexts. The low systematic biases between measured and estimated value indicate a very good agreement between both methods to determine Hbmass and PV. Because PV changes are not considered in the [Hb] thresholds established by the WHO to diagnose anemia, this diagnostic model presents substantial limitations (16, 17). Numerous studies have soundly measured reference hematological values, including [Hb], in populations with high anemia prevalence according to the WHO website, notably in women of reproductive age and young children (< 5 years old) (39-46). Such countries are described with a severe anemia prevalence by the WHO with values going from 40% to 69% in either populations (12, 47). Despite presenting crucial data regarding anemia epidemiology, those studies do not provide any information on either Hbmass or PV, leading to possible over- or underestimation of total prevalence due to PV shifts. Estimating Hbmass and PV with the model presented here could thus, easily and inexpensively produce relevant additional data to assess the severity of anemic cases. Such predictions could increase the sensitivity and specificity of anemia detection by raising the false negative identification and false positive exclusion.

Additionally, strict [Hb] thresholds might not be adapted to all populations. For instance, Otto, Plumb (21) demonstrated the misdiagnosis of anemia in chronic heart failure (CHF) and chronic liver disease (CLD) patients. In those cases, Hbmass remained in healthy ranges, but the diseases induced large PV increases. Such findings are of real clinical importance as substantially low [Hb] triggers additional investigations or treatments whereas hemodilution is often not considered as the potential cause (21). Estimating Hbmass and PV in such cases might thus, effectively decrease the rate of false positive by providing a simple screening tool. Such prediction can also guide clinicians’ decision relative to PBM and transfusion by increasing the robustness of CBC (9). Indeed, it is highly important to correctly assess RBC before engaging in blood transfusion to secure the patients’ health and preserve the transfused blood (8, 10).

Moreover, WHO’ s [Hb] thresholds have also been criticized regarding populations living at altitude. The current guidelines suggest adding from 0.2 g/dL for people living at 1000m to 4.5 g/dL for people living at or over 4500m to the current population cutoffs to account for the increase in RBC synthesis due to the hypoxic environment (11). However, this practice does not relate accurately to the specific population patterns of adaptations to high altitude. It has been shown that Andeans population living at extremely high altitude such as the Aymaras present exceptionally high mean [Hb] (19.1 g/dL), whereas Tibetans and Ethiopians living at similar altitudes have far lower mean values (15.8 g/dL and 15.6 g/dL, respectively) (48, 49). This clearly indicates divergent functional adaptations to similar highly hypoxic environments in those populations and led to calls for specific population thresholds (50-52). Although, those calls are legitimate, they are hard to implement. Moreover, [Hb] is an important phenotype of functional adaptations in populations living at high altitude as demonstrated in Tibetans and Andeans (53). Providing accurate Hbmass and PV estimation would thus, support anemia diagnosis in high altitude populations and help discern between hemoglobin synthesis and volumetric adaptations (54).

Apart from the potential utilization in a diagnostic and clinical context, an accurate prediction of PV can prove to be useful to detect blood doping in sport. The model presented here explained 73% of measured PV variance for women and 79% for men which indicates its potential for individual and longitudinal PV monitoring and could provide complementary data to the Athlete Biological Passport (ABP) hematological module. The ABP hematological module is a tool that longitudinally monitors individual hematological parameters and detects doping by screening non-physiological variations (55, 56). [Hb] being a primary biomarker, the ABP hematological module is subject to multiple confounding factors impacting PV such as acute exercise, heat or altitude exposure (57). Therefore, the PV prediction presented here could provide relevant additional information for the experts reviewing and interpreting those passports (58). Another model estimating PV already exists but requires serum analyses (59). Such analyses present large additional costs and are impossible to implement in large scale longitudinal monitoring. Conversely, estimating PV based on CBC does not imply any supplementary analysis or additional costs.

However, the model introduced here presents some limitations, especially regarding its use in a medical context. Indeed, it was trained on data obtained from healthy and physically active subjects and it questions its application on unhealthy populations. There is thus, a need for data acquired in more comprehensive cohorts to validate the model for medical purposes.

Nevertheless, estimation of Hbmass and PV based on CBC and anthropologic analyses offers highly relevant complementary data about hematological parameters which can prove to be valuable in a wide variety of contexts, from the diagnosis of widespread diseases to anti-doping. It also presents the convenient advantages of being inexpensive, timesaving, and easy to implement.

## Data Availability

The data underlying this article will be shared on reasonable request to the corresponding author.

## Declarations

### Ethics approval and consent to participate

Procedures and risks were fully explained to the subjects, and all of them gave their written consent to participate in the study. All studies were approved by the local ethics committee (CER-VD, Lausanne, Switzerland) and conducted in accordance with the Declaration of Helsinki.

### Consent for publication

Not applicable

### Availability of data and materials

The data underlying this article will be shared on reasonable request to the corresponding author.

### Authors’ contributions

RF designed the study. BM, BK, JS and RF contributed to data collection. BM and JMV performed the regression analyses. BM drafted the first version of the manuscript, and all authors revised it critically. All authors read and approved the final version of the manuscript.

## Acknowledgements

The authors wish to acknowledge Tiffany Rapillard for her help with data collection and all the participants for their participation.

